# Pattern of relapse following three-field lymphadenectomy of esophageal carcinoma and related factors predictive of recurrence

**DOI:** 10.1101/2022.06.20.22276551

**Authors:** Zhi-Chen Xu

**Affiliations:** First Hospital of Quanzhou Affiliated to Fujian Medical University, Quanzhou(362000), People’s Republic of China

**Keywords:** Esophageal cancer, lymph node metastasis, surgery, postoperative radiotherapy

## Abstract

**Background:** For treatment of esophageal carcinoma, the optimal postoperative radiotherapy target volume after three-field lymph node dissection(3-FLD) had not been determined. We analyzed local recurrence pattern of thoracic esophageal carcinoma and risk factors of lymph node recurrence after 3-FLD without prophylactic radiotherapy.

**Methods:** We reviewed 1282 patients with thoracic esophageal squamous cell carcinoma(ESCC) who were treated with 3-FLD without radiotherapy from 2010 to 2018 and analysed local recurrence patterns and risk factors of lymph node recurrence, in order to provide a reference for determination of the radiotherapy target volume for thoracic ESCC.

**Results:** The lymph node recurrence accounted for 91.0% of treatment failures. The mediastinal, cervical and abdominal lymph node recurrence rates were 85.2%, 36.5% and 22.4%, respectively,(x^2^=264.596, P=0.000). The superior, middle and inferior mediastinal lymph node recurrence rates were 67.54%, 27.87% and 0.98%, respectively(x^2^=313.600, P=0.000). In a multivariate analysis, Cevical metastases were significantly associated with N stage and Preoperative cevical lymph node status. Abdominal metastases were significantly associated with the number of preoperative abdominal lymph node metastases(LNM), tumor location and N stage.

**Conclusions:** The main pattern of local-regional recurrence might be lymph node metastasis after radical 3-FLD without radiotherapy in esophageal carcinoma. The dangerous lymph node recurrence regions included neck, superior and middle mediastinum. Radiologist might took the number of pre-operative abdominal lymph nodes and tumor location into consideration while delineating the target area of abdominal region.

This study was supported by Health Department Innovation Fundation Project of Fujian Province (No. 2016-CX-10)

## Introduction

At present, the overall 5-year survival rate of esophageal carcinoma was poor [1]. Despite recent advances in minimally invasive oesophagectomy[2], there was no significant improvement in survival. The main treatment failure causes after surgery were local recurrence and distant metastasis. The local recurrence rates after surgery ranged from 42.9% to 57.9% [3-7]. Although there was no common consensus for postoperative radiotherapy, a growing number of researchers[8-12] suggested that postoperative radiotherapy could reduce local recurrence and was associated with better survival rates for patients with node-positive thoracic esophageal carcinoma (TEC). However, the universally accepted prophylactic radiotherapy target volume for TEC had not yet been established. This study analysed local regional recurrence pattern of thoracic esophageal carcinoma and risk factors of lymph node recurrence after three-field lymph node dissection (3-FLD) without prophylactic radiotherapy, so as to provide evidence for the delineation of the postoperative radiotherapy target volume.

## Methods and Materials

### Patients

From January 2010 to December 2018, 1282 patients with thoracic esophageal squamous cell carcinoma(ESCC) were treated with complete three-field lymph node dissection in our hospital. 335 patients had local-regional recurrence. Among the 335 patients, there were 190 male patients and 145 female patients. Ages ranged from 38 to 77 years, with a median age of 57.5 years old. 101 patients had upper TEC, 176 had middle TEC, and 58 had lower TEC. Patients were staged according to TNM classification (AJCC / UICC, 2009). All patients did not receive preoperative or postoperative radiotherapy. 955 cases received preoperative or postoperative chemotherapy. Patients with palliative resection or apparent residual tumors were excluded. The study protocol was approved by our hospital, and all participants provided written informed consent. The characteristics of the patients were shown in Table 1. Authors had access to information that could identify individual participants during or after data collection.

**Table 1:** Chracteristics of the 1282 patients with thoracic esophageal carcinoma.

| Charateristics | Recurrence | Non-recurrence | $\chi^2$-Value | p-Value |
| --- | --- | --- | --- | --- |
| Gender |  |  |  |  |
| Male | 190 | 538 | 0.001 | 0.976 |
| Female | 145 | 409 |  |  |
| Age/year |  |  |  |  |
| < 60 | 140 | 402 | 0.044 | 0.834 |
| ≥60 | 195 | 545 |  |  |
| Vascular invasion |  |  |  |  |
| Yes | 86 | 221 | 0.741 | 0.389 |
| No | 249 | 726 |  |  |
| T stage |  |  |  |  |
| T1 , T2 | 207 | 669 | 8.963 | 0.003 |
| T3 , T4 | 128 | 278 |  |  |
| N stage |  |  |  |  |
| N0 | 51 | 496 | 173.728 | 0.000 |
| N1 | 103 | 232 |  |  |
| N2 | 130 | 187 |  |  |
| N3 | 51 | 32 |  |  |
| Differentiation |  |  |  |  |
| Poor | 85 | 274 | 2.555 | 0.279 |
| Moderate | 159 | 404 |  |  |
| Well | 91 | 269 |  |  |
| Tumor location |  |  |  |  |
| Upper TEC | 101 | 275 | 1.983 | 0.371 |
| Middle TEC | 176 | 534 |  |  |
| Lower TEC | 58 | 138 |  |  |
| Chemotherapy |  |  |  |  |
| Yes | 221 | 734 | 17.337 | 0.000 |
| No | 114 | 213 |  |  |

Abbreviations: TEC, thoracic esophageal cancer;

### Surgical approach

All patients underwent 3-LND through a right thoractomy, followed by laparotomy, bilateral cervical collar incision. In the thorax, esophagus and the thoracic lymphatic duct were resected. And then the middle and lower mediastinal nodes were removed, which contained the peri-oesophageal, parahiatal, subcarinal and aortopulmonary window nodes. In the abdomen, the upper abdominal and retroperitoneal lymph nodes were removed, which contained coeliac, splenic, common hepatic, left gastric, lesser curvature and parahiatal nodes. In the neck, superior mediastinal and cervical lymph nodes were removed, which contained bilateral recurrent nerve, tracheobronchial, the upper thoracic para-oesophageal nodes, bilateral supraclavicular and deep cervical nodes. More than 15 lymph nodes were dissected during the surgery.

### Follow up

All patients were followed up every 3–6 months after surgery and re-examinations included chest and abdominal enhanced CT, cervical and abdominal Doppler ultrasound and if necessary, PETCT and endoscopy. Positive lymph node should be confirmed by fine needle aspiration cytology. The study was completed on December 31,2021.

### Diagnostic criteria

Preoperative lymph node metastasis was confirmed pathologically after 3-LND. Recurrent diagnosis was mainly based on comprehensive analysis of clinical symptom, physical examination and imaging examination. Local-regional recurrence included lymph nodes recurrence,anastomosis recurrence and tumor bed recurrence. As one criterion, detected nodes were considered recurrent tumors if the long axis was greater than 1 cm [13,14]. In addition, we also took node shape (round or flat), uptake of contrast medium or FDG and difference from the previous node into consideration when evaluating for recurrence. Anastomotic recurrence was determined by histologic analyses following biopsy. Tumor bed recurrence was diagnosed by CT or PET-CT. Any additional recurrences, including other lymph nodes or organs found within 1 month after the initial detection of recurrent tumors, were considered to have occurred simultaneously.

### Terminology of the regional lymph nodes in esophageal cancer

According to a lymph node mapping system for esophageal carcinoma (AJCC-UICC in 2009),The fields of lymph nodes were as follows: 1R(right highest mediastinal nodes); 1L(left highest mediastinal nodes); 2R(right upper paratracheal nodes); 2L(left upper paratracheal nodes); 3P(posterior mediastinal nodes); 4R(right lower paratracheal nodes); 4L(left lower paratracheal nodes); 5(subaortic nodes); 6(paraaortic nodes); 7(subcarinal nodes); 8M(middle paraesophageal lymph nodes); 8L(Lower paraesophageal lymph nodes); 9(pulmonary ligament nodes); 10R(Right hilar nodes); 10L(Left hilar nodes). The lymph nodes in the 1,2,3,4,5 and 6 level were included in the superior mediastinal lymph node; the lymph nodes in the 7, 8M, 10 level were included in the middle mediastinal lymph node; the lymph nodes in the 8L, 9 level were included in the inferior mediastinal lymph node.

### Statistical analysis

SPSS 19.0 statistical software was used for data analysis. A chi square test was used for enumeration data. The predictive factors of recurrence was evaluated by univariate analysis, and logistic regression was used for multivariate analysis. A P value less than 0.05 was considered statistically significant.

## Results

### 1. Recurrent time and pattern

335 patients presented with locoregional recurrence at 8-23 months after surgery, with a median of 14.6 months. The lymph node metastasis, anastomosis recurrence and tumor bed recurrence accounted for 91.0%, 11.9%, and 6.86% of treatment failures, respectively, (x^2^=643.250,P=0.000). The lymph node metastasis was the most common recurrence pattern.

### 2. Lymph node recurrence

Among 305 patients with lymph node recurrence, The mediastinal, cervical and abdominal recurrence accounted for 85.2%, 36.5% and 22.4%, respectively, (x^2^=264.596, P=0.000). There was no significant difference among different segments of the primary tumor in cervical or mediastinal lymph node recurrence. However, The lower TEC had higher metastasis rate of abdominal lymph nodes, compared with upper and middle thoracic esophageal cancer. (x^2^=16.184, P=0.000) (Table 2). The superior, middle and inferior mediastinal lymph node recurrence accounted for 67.54%, 27.87% and 0.98%, respectively,(x^2^=313.600,P=0.000). For superior mediastinal lymph nodes, the station 4R lymph nodes had the highest metastasis rates(29.84%), and there was very significant difference among these regions (x^2^=227.679,P=0.000). For middle mediastinal lymph node, the station 7 lymph nodes had the highest metastasis rates(22.95%), and there was significant difference among these regions (x^2^=92.570, P=0.000). For inferior mediastinal lymph node, locoregional recurrence was rare (Table 3).

**Table 2.**
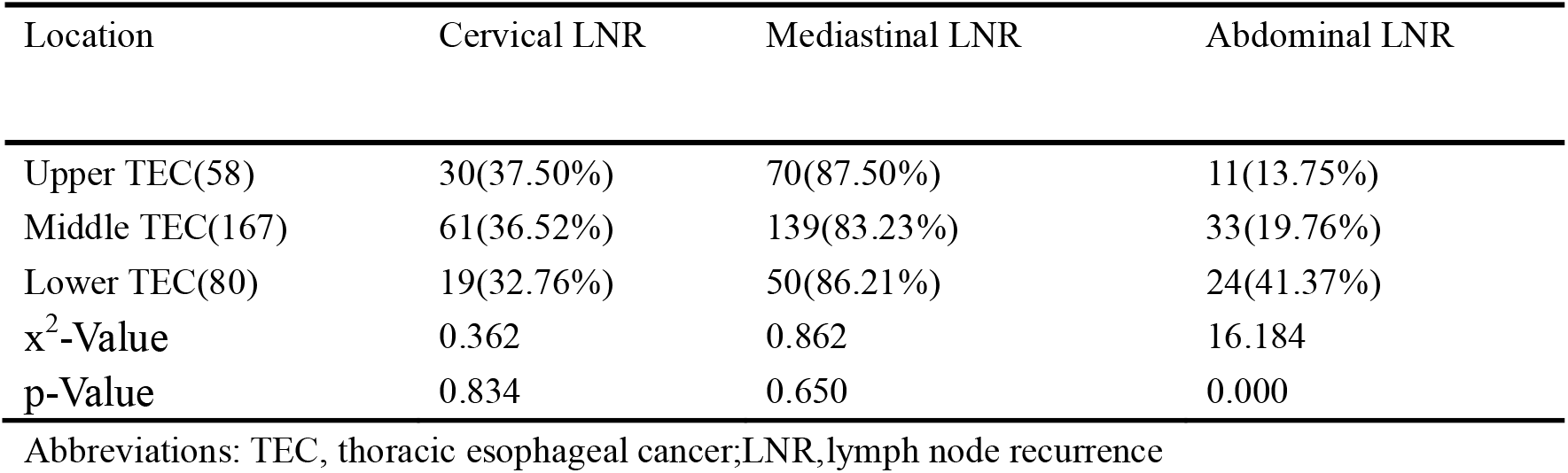
The distribution of lymph node recurrence in patients with thoracic esophageal carcinoma.

| Location | Cervical LNR | Mediastinal LNR | Abdominal LNR |
| --- | --- | --- | --- |
| Upper TEC(58) | 30(37.50%) | 70(87.50%) | 11(13.75%) |
| Middle TEC(167) | 61(36.52%) | 139(83.23%) | 33(19.76%) |
| Lower TEC(80) | 19(32.76%) | 50(86.21%) | 24(41.37%) |
| $\chi^2$-Value | 0.362 | 0.862 | 16.184 |
| p-Value | 0.834 | 0.650 | 0.000 |

Abbreviations: TEC, thoracic esophageal cancer; LNR, lymph node recurrence

**Table 3.** The distribution of mediastinal lymph node recurrence in patients with thoracic esophageal carcinoma.

| Site of LNM | recurrence(%) | Site of LNM | recurrence(%) |
| --- | --- | --- | --- |
| SM LNM | 206(67.54%) | Station 6 | 7(2.30%) |
| Station 1R | 58(19.02%) | MMLNM | 85(27.87%) |
| Station 2R | 51(16.72%) | Station 7 | 70(22.95%) |
| Station 4R | 91(29.84%) | Station 8M | 14(4.59%) |
| Station 1L | 45(14.75%) | Station 10R | 15(4.92%) |
| Station 2L | 40(13.11%) | Station 10L | 13(4.26%) |
| Station 4L | 61(20.00%) | IMLNM | 3(0.98%) |
| Station 5 | 67(18.69%) | Station 8L | 3(0.98%) |
| Station 3A | 0(0.00%) | Station 9 | 0(0.00%) |
| Station 3P | 0(0.00%) |  |  |

Abbreviations: SMLNM: Superior mediastinal lymph node metastasis; MMLNM: Middle mediastinal lymph node metastasis; IMLNM: Inferior mediastinal lymph node metastasis
SMLNM: $\chi^2=227.679$ , $P=0.000$ ; MMLNM: $\chi^2=92.570$ , $P=0.000$ ;

### 3. Characteristics and risk factors of cervical recurrence

Among 110 patients with cervical recurrence, right supraclavicular, left supraclavicular, right cervical paraesophageal and left cervical paraesophageal lymph nodes recurrence accounted for 48.18%, 25.45%, 18.18%, and 13.63%, respectively (x^2^=39.991, P=0.000), while the rest of abdominal lymph node recurrence were rare. The cervical lymph node recurrence in the right was significantly higher than that in the left (x^2^=16.412, P=0.000). Univariate analysis revealed no significant differences in cervical recurrence in terms of gender, age, vascular invasion, T stage, differentiation, tumor location, chemotherapy. However, significant differences were observed in relation to N stage (x^2^=113.671, P=0.000) and Preoperative cevical lymph node status (x^2^=27.981, P=0.000). Then we analyzed the above two clinical factors and cevical lymph node recurrence by multivariate regression analysis. The results indicated that cevical metastases were significantly associated with N stage and preoperative cevical lymph node status (Table 4). In patients with postoperative pathology of N 0, 1, 2, 3, the cevical recurrence rates were 2.0%, 7.2%, 14.8% and 33.7%, respectively. The cevical metastasis rates of negative and positive cevical lymph nodes were 2.6% and 11.5%, respectively.

**Table 4:** Multiple logistic regression analysis of risk factors of cervical lymph node recurrence in patients with thoracic esophageal carcinoma.

| Variable | Regression coefficient B | S.E | Wald | HR(95%CI) | P-Value |
| --- | --- | --- | --- | --- | --- |
| N stage | 0.952 | 0.115 | 68.118 | 2.592(2.067-3.250) | 0.000 |
| Preoperative cervical LNM | 1.125 | 0.333 | 11.436 | 3.081(1.605-5.915) | 0.001 |

Abbreviations: LNM, lymph node metastases

### 4. Characteristics and risk factors of abdominal recurrence

Among 68 patients with abdominal recurrence, lymph node recurrence at the para-aortic(above the left renal vein), celiac artery, posterior surface of the pancreatic head, and common hepatic artery accounted for 77.94%, 44.11%, 27.94%, and 23.52%, respectively(x^2^=50.592, P=0.000), while the rest of abdominal lymph node recurrence were rare. Univariate analysis revealed no significant differences in abdominal recurrence in terms of gender, age, vascular invasion, T stage, differentiation and chemotherapy. However, significant differences were observed in relation to the number of preoperative abdominal lymph node metastases(LNM) (x^2^=99.870, P=0.000), tumor location (x^2^=23.642, P=0.000) and N stage (x^2^=62.665, P=0.000). In a multivariate analysis, abdominal metastases were significantly associated with the number of preoperative abdominal LNM, tumor location and N stage(Table 5). The abdominal metastasis rates were 2.0% for those with 0 positive abdominal nodes, 11.4% for 1 to 2 nodes, and 20.0% for ≥3 nodes, respectively. The rates of abdominal metastasis from upper, middle, and lower esophageal thoracic cancers were 2.9%, 4.6%, and 12.2%, respectively. In patients with pathology of N0,1,2,3, the abdominal recurrence rates were 0.7%, 4.5%,11.7% and 14.5%, respectively.

**Table 5:** Multiple logistic regression analysis of risk factors of abdominal Lymph node recurrence in patients with thoracic esophageal carcinoma.

| Variable | Regression coefficient B | S.E | Wald | HR(95%CI) | P-Value |
| --- | --- | --- | --- | --- | --- |
| No of Preoperative abdominal LNM | 0.980 | 0.173 | 31.960 | 2.665(1.897-3.744) | 0.000 |
| Tumor location | 0.870 | 0.213 | 16.606 | 2.386(1.570-3.625) | 0.000 |
| N stage | 0.527 | 0.178 | 8.804 | 1.694(1.196-2.399) | 0.003 |

Abbreviations: LNM, lymph node metastases

## Discussion

Surgery remained the standard treatment for TEC. However, the optimal postoperative radiotherapy target volume for treatment of esophageal carcinoma had not been determined. The delineation of postoperative radiation target volume in many centers was mostly based on the pattern of lymphatic spread[15]. The treatment area included bilateral supraclavicular area, mediastinum and abdominal lymph nodes. This target volume was too large and patients frequently suffer from complications such as gastrointestinal reactions, radiation pneumonitis, esophagitis and leucopenia. Furthermore, this target volume might be inconsistent with recurrence pattern and result in missing the target area. Fujita et al. [16] reported that in patients with upper TEC, metastasis at operation was commonly found in the cervicothoracic and periesophageal nodes, and recurrence mostly occurred in the cervical nodes; in patients with lower TEC, metastasis at operation was commonly found in the periesophageal nodes and upper abdominal nodes, whereas recurrence mostly occurred in the celiac nodes. An irradiation field that is too large would cause unnecessary radiation damage, whereas an overly small field would be miss cancerous cells. Thus, we analyzed the recurrence patterns and risk factors in esophageal carcinoma patients after 3-FLD to provide a reference for the determination of the optimal target volume for postoperative radiotherapy.

In this study, the lymph node recurrence was the most common local regional recurrence. The common lymph node recurrence regions were neck, superior and middle mediastinum. This result was consistent with some studies[17-20]. A possible reason for this result was the technical difficulty of lymph node dissection in the superior, and middle mediastinal regions. This might be due to complex anatomy with abundant nerves, lymphatic vessels, and adjoining large blood vessels and critical organs in these regions. A few residual cancer cells, particularly in advanced TEC, might lead to recurrence after surgery. On the contrary, the inferior mediastinal and upper abdominal areas could be well exposed, and lymph node dissection was comparatively more thorough. Further research indicated that mediastinal recurrence was not significantly affected by vertical location. Therefore, we suggested the postoperative irradiation of station 1–5 and 7 lymph nodes should be carried out prophylactically for all TEC segments.

To date, whether the cervical region should be included in the postoperative target volume remained controversial. Lu et al.[21] reported that after surgery, supraclavicular lymph node metastasis in patients in the lower and middle lower thirds were 1/43 and 1/18. So, it seems unnecessary that the bilateral supraclavicular area should be irradiated in the lower and middle lower thirds. In this study, the recurrence rate of cervical lymph node was 36.5%, consistent with Cai et al reports[20]. Further analysis indicated that cevical metastases were significantly associated with N stage and Preoperative cevical lymph node status. Therefore, our research suggested that cervical areas, including the supraclavicular and cervical paraesophageal lymph node, should be included in the postoperative radiotherapy target volume in consideration of its high recurrence rate, especially for patients with preoperative cervical lymph node metastasis.

Whether to irradiate the abdominal area remained controversial. In this study, we determined abdominal metastases rate of 20.29% (68/335), consistent with previous study[20,22]. Multivariate analysis indicated that abdominal metastases were significantly associated with tumor location, the number of preoperative abdominal LNM and N stage. The abdominal metastasis rate of lower TEC(12.2%) was significantly higher than that for the upper(2.9%) and middle(4.6%) TECs. This result was consistent with previous studies[23-25]. This might be attributed to that lower TEC might mainly metastasize to abdominal para-aortic lymph nodes. The abdominal metastasis rates were 2.0% for patients with 0 positive abdominal nodes, 11.4% for 1 to 2 nodes, and 20.0% for ≥3 nodes, respectively. These findings indicated that the number of abdominal LNM at operation might be an important predictor of abdominal lymph node recurrence. This could be attributed to the technically difficulty in removing all lymph nodes adjacent to the abdominal aorta, celiac artery, posterior surface of the pancreatic head, and common hepatic artery. These few residual cancer cells might result in recurrence[24]. Therefore, we suggested these abdominal areas should be irradiated for lower TEC patients with preoperative abdominal LNM.

One of the shortcomings of this study was that this study was a retrospective analysis, a clear conclusion could not be exactly drawn. A large sample prospective study with postoperative radiotherapy followed by 3-LND in patients with high risk factors seemed necessary to assess the postoperative radiation value for local control.

In conclusion, The main pattern of local-regional recurrence of esophageal squamous cell carcinoma after radical 3-FLD without prophylactic radiotherapy might be lymph node metastasis. Common metastasis regions were cervical, superior and middle mediastinal regions. Cevical recurrence was significantly associated with N stage and preoperative cevical LNM. Abdominal recurrence was significantly associated with number of preoperative abdominal LNM, tumor location and N stage. We regarded that Radiologist might took the number of pre-operative abdominal lymph nodes and tumor location into consideration while delineating the target area of abdominal region.

## Data Availability

Data cannot be shared publicly .

## Ethic Approval

Approval to undertake this study was sought and granted by the Ethic Committee of Quanzhou First Hospital.

